# Electrocardiogram-Based Deep Learning for Time-Resolved Prediction of Heart Failure With Reduced Ejection Fraction: A Multinational Study

**DOI:** 10.64898/2026.07.08.26356558

**Authors:** Lei Pan, Siyuan Li, Jiayu Huo, Zilong Xiao, Ziqing Yu, Juecheng Chen, You Zhou, Zhixing Li, Beijian Zhang, Xiao Li, Cong Wang, Hongyang Lu, Konstantinos Patlatzoglou, Daniel B. Kramer, Jonathan W. Waks, Fu Siong Ng, Yixiu Liang, Junbo Ge

## Abstract

**Background:** Heart failure with reduced ejection fraction (HFrEF) remains a major global health burden. Most electrocardiogram (ECG)-based artificial intelligence models are limited to diagnostic tasks or fixed-horizon prognostic classification and provide little insight into the temporal evolution of risk. In addition, concerns regarding model interpretability continue to impede clinical adoption. Whether deep learning applied to ECGs can deliver individualized, time-resolved, and biologically interpretable risk estimates for incident HFrEF across diverse populations remains uncertain.

**Methods:** We developed a convolutional neural network-based survival model using raw 12-lead ECGs from Zhongshan Hospital (SHZS) and externally validated it in independent cohorts from Shanghai Tenth People’s Hospital (SHTP) and Beth Israel Deaconess Medical Center (BIDMC). The model generated individualized, day-by-day probabilities of incident HFrEF over a 5-year horizon. Performance was comprehensively evaluated using discrimination, calibration, precision-recall characteristics, clinical utility, and risk stratification metrics, with subgroup analyses across age, sex, and race to assess generalizability. Model interpretability was examined using complementary representation and attention-based frameworks.

**Results:** In 458,884 patients, the survival model demonstrated strong and stable discrimination across cohorts, with overall C-indices of 0.971 (95% CI, 0.965-0.976) in SHZS, 0.945 (95% CI, 0.938-0.950) in SHTP, and 0.855 (95% CI, 0.850-0.860) in BIDMC, and consistently high time-dependent AUROC values across the 1-5-year horizons. Calibration showed close agreement between predicted and observed risks, and decision curve analyses indicated meaningful net clinical benefit across a broad range of thresholds. Kaplan-Meier curves showed clear stratification across predicted risk groups. Interpretability analyses identified physiologically coherent ECG features related to QRS duration, heart rate, and QT interval that were associated with predicted risk.

**Conclusion:** This ECG-based deep learning survival model provides individualized, time-resolved, and clinically interpretable estimates of future HFrEF risk with robust performance across multinational cohorts. These findings support the potential of AI-enabled ECG analysis as an accessible tool for early HFrEF risk stratification within routine clinical workflows.

## Introduction

Heart failure (HF) with reduced ejection fraction (HFrEF) accounts for nearly half of all HF hospitalizations and is associated with an approximately 50% mortality within five years of diagnosis. **^[1]^**Despite the availability of effective therapies, delayed recognition of left ventricular systolic dysfunction remains common in routine clinical care, particularly when early symptoms are absent or nonspecific.**^[2]^** Early identification of individuals at risk could enable timely intervention and improve long-term outcomes.**^[3–6]^** Although biomarkers such as N-terminal pro-B-type natriuretic peptide (NT-proBNP) and high-sensitivity cardiac troponin are independently associated with incident heart failure,**^[7–10]^** their clinical application is limited by the need for invasive testing and poor accessibility in routine care. **^[11]^**Likewise, clinical risk scores require specialized assessments and exhibit variable predictive accuracy and feasibility in real-world settings. **^[12–14]^**There remains, therefore, a critical unmet need for inexpensive and accessible prognostic tools to identify HFrEF patients.

Artificial intelligence-enabled electrocardiogram (AI-ECG) systems have emerged as promising tools for predicting HF outcomes from a single examination**.^[15]^** However, most existing models remain confined to diagnostic classification or prognostic frameworks that provide only static or binary risk estimates, without capturing the temporal evolution of risk. **^[2,16–20]^** Importantly, this limitation substantially reduces their clinical utility, as identifying when a patient is most vulnerable is often more informative than determining whether risk is present. In addition, reported AI-ECG performance frequently varies across age, sex, and race, raising concerns regarding generalizability and hindering broader clinical implementation.**^[21]^**

We hypothesized that AI-ECG could not only identify individuals at risk of heart failure but also estimate individualized time-to-event probabilities of disease onset, thereby capturing the temporal evolution of risk rather than relying on static classification. Such models could support earlier identification and timely follow-up of vulnerable patients within clinical workflows. To test this hypothesis, we developed and evaluated a survival-based deep learning model trained on ECG data, with validation across geographically and ethnically diverse populations to assess generalizability across key demographic groups.

## Methods

### Study population

The derivation cohort was obtained from Zhongshan Hospital, Shanghai (SHZS), and included all individuals aged ≥14 years, corresponding to the age of adult assessment at this center, who underwent at least one 12-lead ECG and transthoracic echocardiogram (TTE) between February 2014 and May 2020. Within this cohort, data were randomly partitioned into training (80%), validation (10%), and internal test (10%) sets for model development and performance assessment.

External testing was performed using two independent datasets. The first external cohort was derived from the Tenth People’s Hospital of Shanghai (SHTP) and included patients who underwent at least one ECG and TTE between April 2017 and March 2023 during routine medical care. The second external cohort was obtained from Beth Israel Deaconess Medical Center (BIDMC) and consisted of individuals aged ≥16 years with valid ECG recordings between 2000 and 2023 in both outpatient and inpatient settings.

Collectively, these cohorts encompassed a heterogeneous population ranging from healthy individuals to patients with a broad spectrum of cardiac and non-cardiac conditions.

The study was approved by the Institutional Research Board of Zhongshan Hospital (No. B2023-253R) with a waiver of patient consent, and registered on ClinicalTrials.gov (NCT07519434). For the BIDMC cohort, ethics review and approval was provided by the Beth Israel Deaconess Medical Center Committee on Clinical Investigations, IRB protocol # 2023P000042. All study procedures were performed in compliance with the Declaration of Helsinki.

### Data collection and pre-processing

Raw ECG data contained eight independent leads stored in the digital acquisition system, from which the remaining leads were computationally derived to reconstruct standard 12-lead ECG signals. The signals were preprocessed using a 0.5-100 Hz bandpass filter combined with a 50 Hz notch filter to remove baseline drift and powerline interference and were subsequently resampled to 500 Hz. To reflect real-world clinical practice, no ECG recordings were excluded because of noise or signal artifacts. Each 10-second ECG recording was zero-padded to 5000 samples per lead and used as input to the deep learning model. For TTE, left ventricular ejection fraction (LVEF) values were extracted from the final echocardiographer diagnostic reports in structured tabular format.

ECG-TTE pairing was performed using a predefined hierarchical strategy. Only patients with at least one TTE performed within 30 days of an ECG were eligible for inclusion. When an ECG corresponded to multiple TTEs with discordant findings, the ECG was paired with the earliest TTE demonstrating LVEF ≤40%; in the absence of positive findings, the TTE obtained at the longest interval was selected to maximize follow-up. For classification model development, patients with negative TTE findings and a follow-up duration of less than five years were excluded.**^[22]^**

### Model development

The endpoint of this study was defined as the occurrence of HFrEF, characterized by a LVEF ≤40% during follow-up. To address this task, we built our models by fine-tuning a ECG Founder backbone on our dataset**.^[23]^** Two complementary modeling strategies were implemented. First, a diagnostic classification model was trained using a binary outcome of HFrEF at baseline, derived from ECGs paired with TTEs obtained within 30 days (n = 20,742 ECG-TTE pairs from 13,679 patients). Second, a prognostic survival model was developed to simultaneously capture prevalent HFrEF and estimate the risk of future onset. Unlike the classification setting, this approach incorporated all available ECG-TTE pairs regardless of temporal distance, with the interval length directly encoded in the model (n = 319,801 ECG-TTE pairs from n = 240,538 patients). The CNN output layer was adapted to a discrete-time survival structure, allowing the model to learn both time-to-event information and censoring patterns. In this framework, baseline HFrEF (LVEF ≤40%) was encoded at the initial time step, while subsequent steps reflected the probability of incident HFrEF over time. Both diagnostic performance and prognostic performance were assessed at a 5-year horizon.

### Model evaluation

Model performance was evaluated using several complementary metrics. Discrimination was quantified with the concordance index (C-index), and the mean area under the receiver operating characteristic curve (AUC) was calculated annually to assess time-dependent predictive ability. To account for outcome imbalance, the area under the precision-recall curve (AUPRC) was also reported. Calibration was examined using annual Brier scores, defined as the average squared error between predicted survival probabilities and observed outcomes. Clinical utility was further explored through decision curve analysis (DCA), which estimates the net benefit across a range of threshold probabilities. For risk stratification, patients were divided into low-, intermediate-, and high-risk groups according to their predicted 3-year probabilities of developing HF with LVEF ≤40%. Kaplan-Meier curves were then constructed to display differences in HF incidence among these risk categories, and statistical significance was evaluated using the log-rank test.

### Model comparison

The performance of the AI-ECG model was compared with the pooled cohort equations to prevent heart failure (PCP-HF), a validated clinical risk model incorporating demographic, clinical, laboratory, medication, and ECG-derived variables. Comparative analyses were restricted to the BIDMC cohort owing to incomplete availability of required PCP-HF input features in the SHZS and SHTP cohorts.

The incremental prognostic value of the proposed model beyond the PCP-HF model was assessed using integrated discrimination improvement (IDI) and both categorical and continuous net reclassification improvement (NRI). These metrics were estimated in a time-dependent manner at prespecified horizons (years 1-5) to evaluate improvements in longitudinal risk stratification.

### Imaging association analysis

To explore the biological plausibility of the AI-ECG predictions, imaging association analyses were performed by examining correlations between AI-ECG-derived predictions and TTE measurements in the BIDMC cohort. Univariate correlations were assessed between AI-ECG predictions and routinely measured TTE parameters.

### Interpretability analysis

To investigate ECG patterns contributing to the model’s predicted risk of developing HFrEF, we applied a variational autoencoder (VAE)-based interpretability framework. Median beats were extracted using the BRAVEHEART software and used to train a convolutional VAE that learned compact latent representations of ECG morphology. Latent features from the encoder were then evaluated using a light gradient boosting machine, with SHAP values used to identify the most influential latent dimensions. For these key features, latent-space interpolation was performed to visualize how variations in each latent factor modified the corresponding ECG waveforms. In addition, correlations between VAE-derived latent features and conventional ECG parameters were computed to generate heatmaps illustrating the physiological relevance of the latent dimensions.

To further probe model-level interpretability, median-beat reconstructions were combined with Gradient-weighted Class Activation Mapping (Grad-CAM). Median-beat signals from 1000 subjects with the lowest and highest predicted HFrEF risks were also averaged and compared to highlight characteristic waveform differences. The resulting importance maps were superimposed onto median-beat waveforms for each lead, enabling simultaneous visualization of both the model’s attention patterns and the morphological differences between high- and low-risk groups.

### Statistical analysis

Continuous variables are summarized as mean ± standard deviation, whereas categorical variables are presented as counts with corresponding percentages. Group comparisons for continuous measures were conducted using Student’s t-test, and categorical data were assessed with the chi-square test. For model evaluation, the Likelihood Ratio test was applied to nested Cox models, while the Partial Likelihood Ratio test was used for comparisons between non-nested models. A two-sided p value < 0.05 was considered indicative of statistical significance. All statistical analyses were carried out using R software (version 4.2.0; R Core Team, Vienna, Austria) and Python (version 3.10.12).

### Code availability

The code for model training and evaluation is publicly available at Github: https://github.com/lsy2006/AIECG-HF. Access to the underlying data is restricted due to patient privacy and institutional regulations.

## Results

### Patient characteristics

A total of 37,891 patients from the SHZS cohort were included as the internal test set. External validation was performed in 143,129 patients from the SHTP cohort and 29,046 patients from the BIDMC cohort. The overall study workflow and patient selection process are illustrated in **Figure 1**.

**Figure 1.**
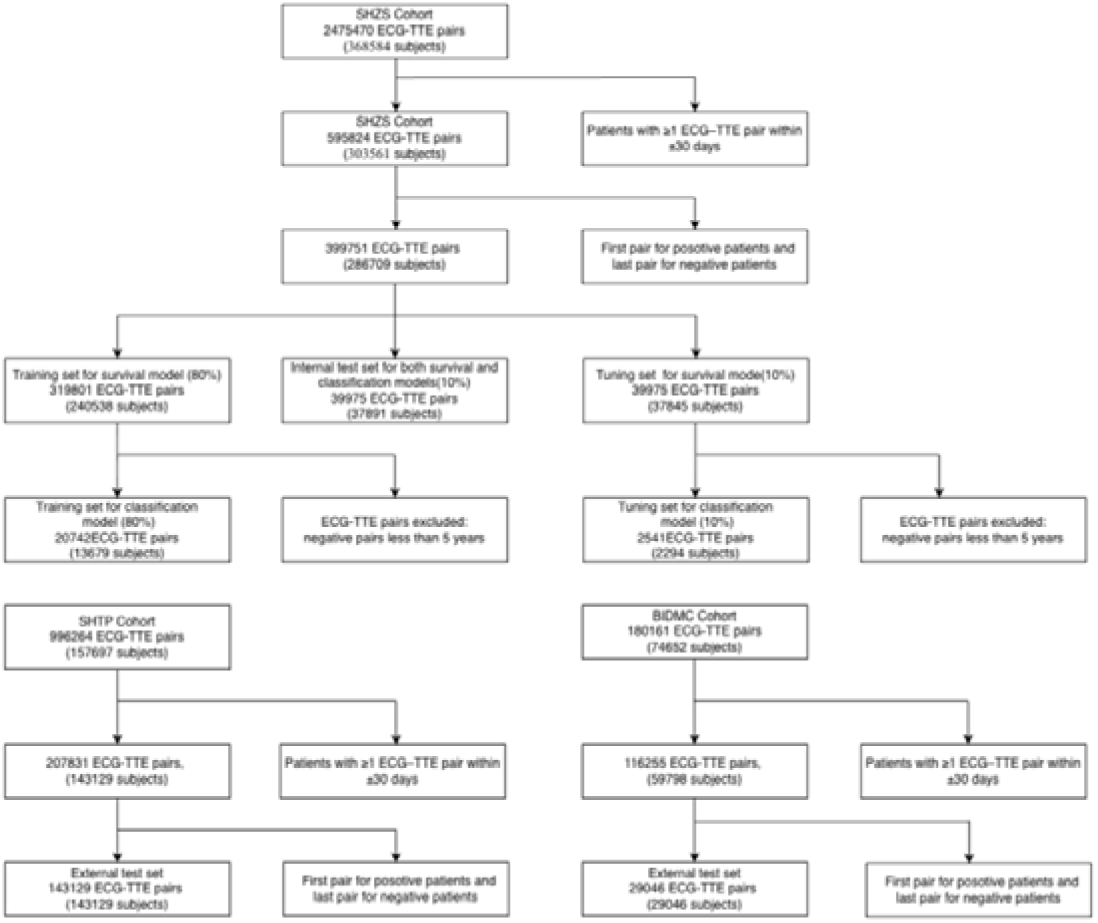
Study flow diagram. Diagram depicts data used to train and evaluate the AI-ECG survival and classification models. ECG, electrocardiogram; TTE, transthoracic echocardiogram; SHZS, Shanghai Zhongshan Hospital; SHTP, Shanghai Tenth People’s Hospital; BIDMC, Beth Israel Deaconess Medical Center.

At baseline, significant heart failure (defined as LVEF ≤40%) was present in 993 (2.48%) ECG-TTE pairs in SHZS, 2,299 (1.61%) in SHTP, and 5,249 (18.07%) in BIDMC, demonstrating marked differences in baseline prevalence across cohorts. Among individuals with preserved baseline LVEF (>40%) who underwent repeat echocardiography ≥30 days after the index ECG, progression to LVEF ≤40% occurred in 150 (0.38%) participants in SHZS, 470 (0.33%) in SHTP, and 2,162 (7.44%) in BIDMC during mean follow-up periods of 1.84, 0.3, and 3.67 years, respectively.

Baseline demographic and clinical characteristics, together with echocardiographic data, are summarized in **Table 1**.

**Table 1.**
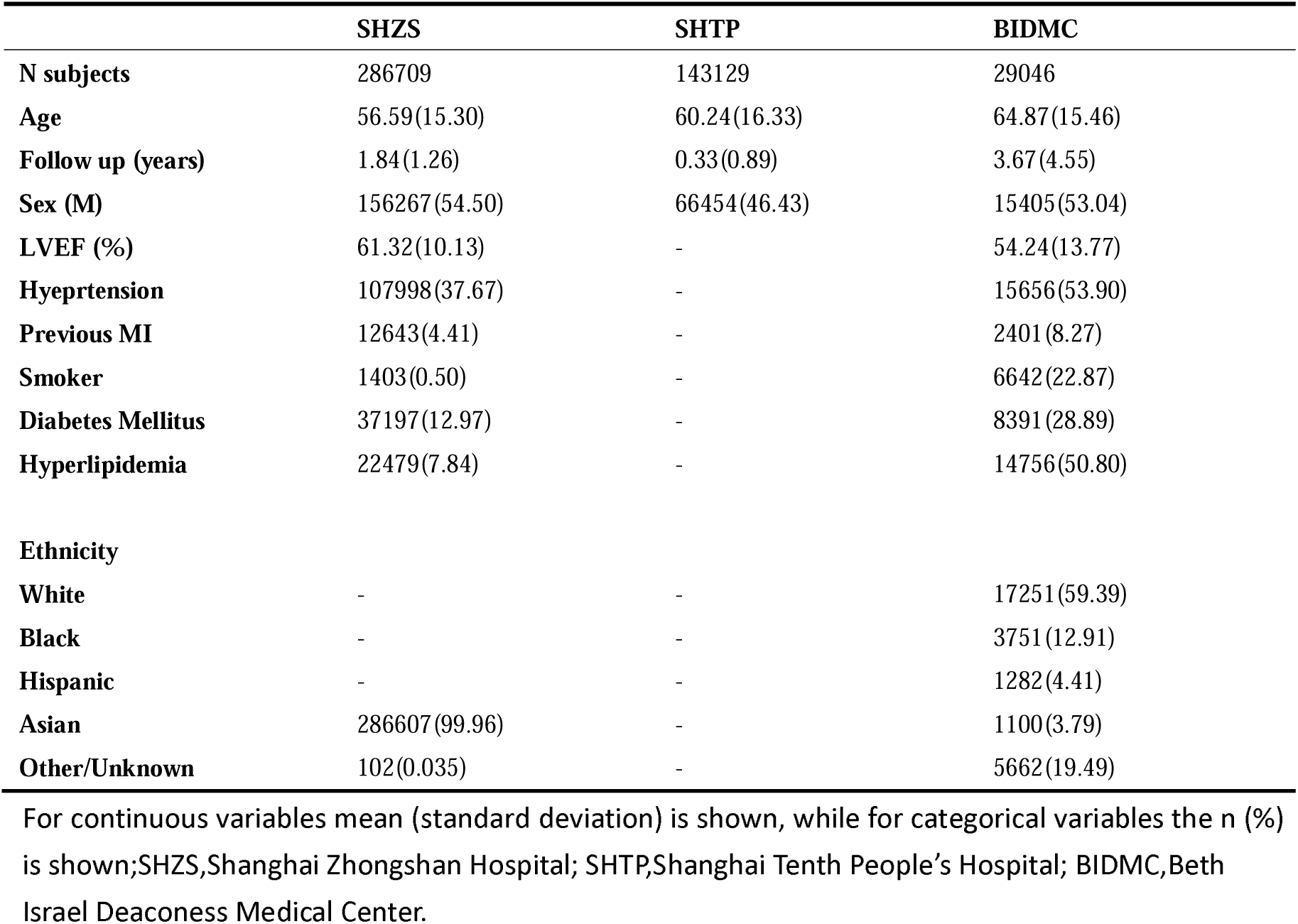
Population Characteristics of the Study Cohorts.

|  | SHZS | SHTP | BIDMC |
| --- | --- | --- | --- |
| **N subjects** | 286709 | 143129 | 29046 |
| **Age** | 56.59(15.30) | 60.24(16.33) | 64.87(15.46) |
| **Follow up (years)** | 1.84(1.26) | 0.33(0.89) | 3.67(4.55) |
| **Sex (M)** | 156267(54.50) | 66454(46.43) | 15405(53.04) |
| **LVEF (%)** | 61.32(10.13) | - | 54.24(13.77) |
| **Hypertension** | 107998(37.67) | - | 15656(53.90) |
| **Previous MI** | 12643(4.41) | - | 2401(8.27) |
| **Smoker** | 1403(0.50) | - | 6642(22.87) |
| **Diabetes Mellitus** | 37197(12.97) | - | 8391(28.89) |
| **Hyperlipidemia** | 22479(7.84) | - | 14756(50.80) |
| **Ethnicity** |  |  |  |
| **White** | - | - | 17251(59.39) |
| **Black** | - | - | 3751(12.91) |
| **Hispanic** | - | - | 1282(4.41) |
| **Asian** | 286607(99.96) | - | 1100(3.79) |
| **Other/Unknown** | 102(0.035) | - | 5662(19.49) |

For continuous variables mean (standard deviation) is shown, while for categorical variables the n (%) is shown; SHZS, Shanghai Zhongshan Hospital; SHTP, Shanghai Tenth People's Hospital; BIDMC, Beth Israel Deaconess Medical Center.

### Discrimination and overall predictive performance across cohorts

Overall discrimination of the survival model was high, with C-indices of 0.971 (95% CI, 0.965–0.976) in SHZS, 0.945 (95% CI, 0.938–0.950) in SHTP, and 0.855 (95% CI, 0.850–0.860) in BIDMC. Across follow-up, the time-dependent C-index remained stable at 0.970–0.972 in SHZS, 0.949–0.950 in SHTP, and 0.852–0.856 in BIDMC, while AUROC remained above 0.96 in SHZS, ranged from 0.922 to 0.935 in SHTP, and was not below 0.869 in BIDMC (**Figure 2A and 2B**).

**Figure 2.**
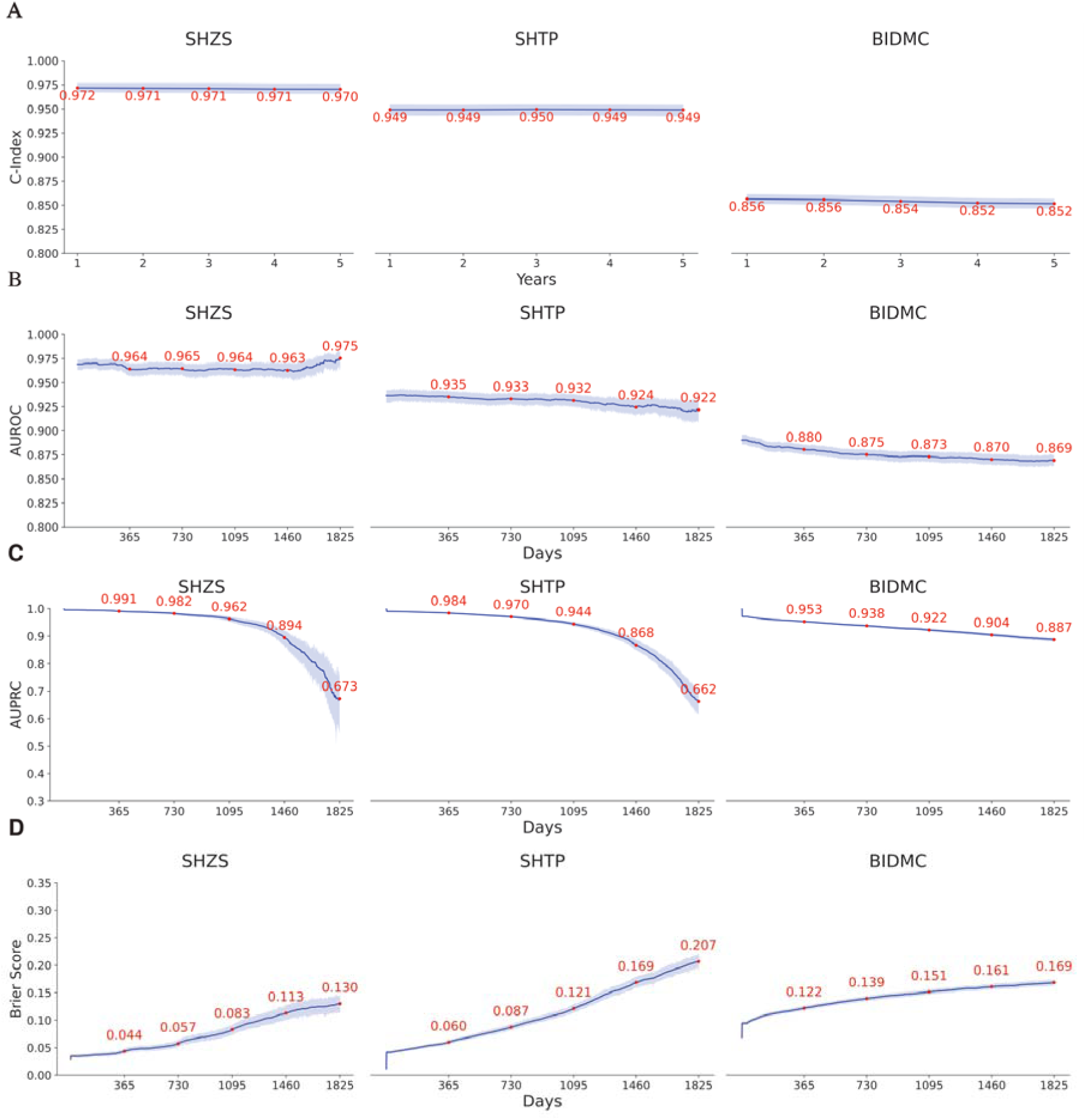
Time-dependent predictive performance of the AI-ECG survival model across cohorts. Model performance was evaluated in the SHZS, SHTP, and BIDMC cohorts using time-dependent metrics. (A) Time-dependent C-index across prediction horizons (Years 1-5). (B) Time-dependent area under the receiver operating characteristic curve (AUROC) over follow-up time. (C) Time-dependent area under the precision-recall curve (AUPRC) over follow-up time. (D) Time-dependent Brier score over follow-up time. Curves represent estimated metric values over time, with shaded bands indicating 95% confidence intervals; red numeric labels denote metric values at prespecified time points.

Precision–recall curves showed the expected gradual decline over longer horizons, with AUPRC decreasing from 0.991 to 0.673 in SHZS and from 0.984 to 0.662 in SHTP, reflecting lower event density at later time points. Notably, AUPRC remained comparatively higher in BIDMC (0.953–0.897), consistent with its higher baseline event rate (**Figure 2C**). Brier scores increased gradually over longer prediction horizons, as expected, but remained relatively low in SHZS (0.044–0.130) and BIDMC (0.122–0.169), with slightly greater increases in SHTP (0.060–0.207), indicating limited error accumulation during longitudinal prediction (**Figure 2D**).

### Calibration performance across cohorts

Consistent with the Brier score findings, calibration analyses showed good agreement between predicted and observed risks across all cohorts (**Figure 3A**). In SHZS, the model showed slight underestimation at lower predicted probabilities and modest overestimation at higher risk levels, with similar minor deviations observed in SHTP and BIDMC. Consistent calibration was observed across the 1-, 2-, 4-, and 5-year horizons (**Figure S1**).

**Figure 3.**
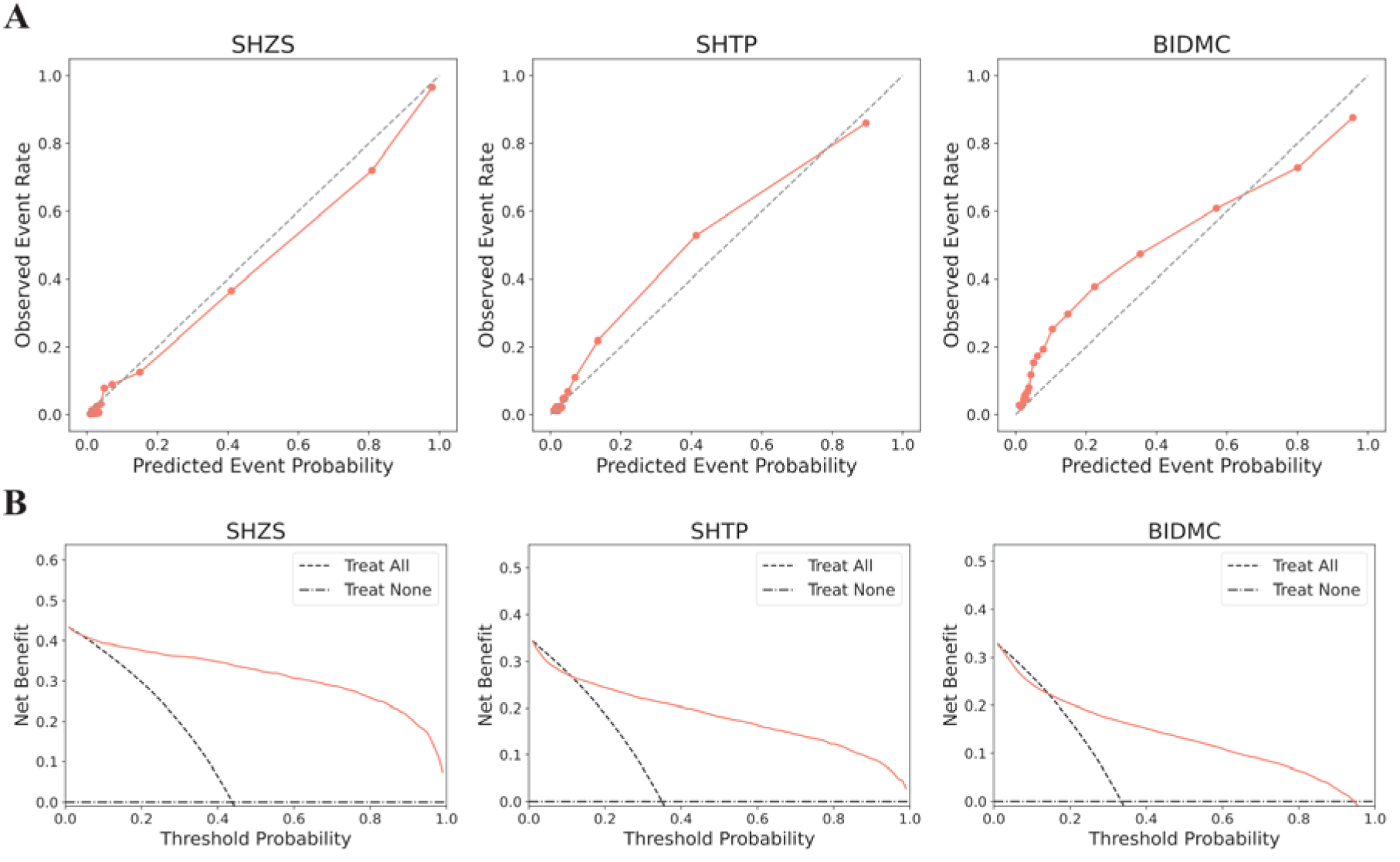
Calibration and clinical utility of the AI-ECG survival model across cohorts at 3 years. Model calibration and clinical utility were evaluated in the SHZS, SHTP, and BIDMC cohorts at the 3-year prediction horizon. (A) Calibration plots comparing predicted event probabilities with observed event rates across risk deciles; the dashed diagonal line indicates perfect calibration. (B) Decision curve analysis (DCA) demonstrating the net clinical benefit of the AI-ECG model across a range of threshold probabilities, compared with treat-all and treat-none strategies.

### Clinical utility analysis

Decision curve analysis showed that the model provided the greatest clinical benefit during the early to mid-term horizons in SHZS and SHTP, with clear net benefit at 1–3 years and attenuated advantage at 4–5 years (**Figure 3B and Figure S2**). In SHZS, benefit at 4 years was retained above a threshold probability of 20%, whereas at 5 years it was limited to very high thresholds; similar patterns were observed in SHTP. By contrast, BIDMC showed a more consistent pattern across the 1- to 5-year horizons, with net benefit generally exceeding the treat-all strategy across most threshold probabilities after a small crossover at lower thresholds.

### Survival analysis across cohorts

In the SHZS internal cohort, patients classified as high risk exhibited a markedly lower survival probability than those in the medium- and low-risk groups at the 3-year horizon (log-rank p < 0.0001). Similar risk stratification patterns were observed in the SHTP and BIDMC external cohorts (**Figure 4**). Consistent separation of survival curves across low-, medium-, and high-risk groups was also evident at 1-, 2-, 4-, and 5-year time points in all three cohorts (**Figure S3)**.

**Figure 4.**
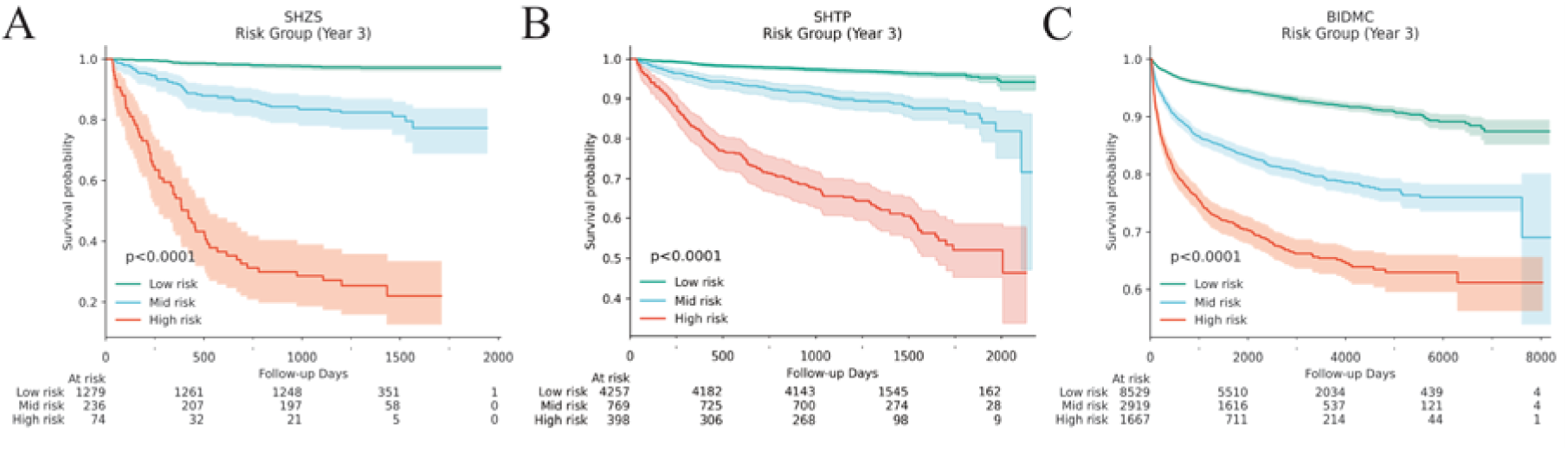
Kaplan-Meier survival curves stratified by AI-ECG-predicted risk at 3 years across cohorts. Kaplan-Meier survival curves in the SHZS, SHTP, and BIDMC cohorts, stratified by AI-ECG-predicted risk groups derived from the 3-year prediction horizon (low, medium, and high). Shaded areas represent 95% confidence intervals. Numbers at risk are shown below each panel. Survival differences among risk groups were assessed using the log-rank test (p < 0.0001 for all cohorts).

### Subgroup analyses

The model maintained strong discrimination across age and sex subgroups in SHZS, SHTP, and BIDMC (**Figure S4**). In SHZS, C-indices ranged from approximately 0.95 to 0.97 and AUROCs from 0.95 to 0.98, whereas in the external cohorts the corresponding ranges were 0.85–0.91 and 0.86–0.93. Race-based subgroup analyses in BIDMC showed similar performance across White, Black, Asian, Hispanic, and other racial groups, with C-indices ranging from approximately 0.85 to 0.88 and AUROC values from 0.86 to 0.93. Additional performance metrics yielded concordant findings across subgroups.

### Performance comparison between survival and classification models

Across all three cohorts, the survival model consistently outperformed the classification model in discrimination and overall predictive accuracy (**Figure S5**). In SHZS and SHTP, the survival model achieved higher C-indices and time-dependent AUROCs across nearly all prediction horizons, with similar superiority observed in BIDMC. AUPRC was also generally higher for the survival model, whereas Brier scores were consistently lower, indicating better probability estimation. These differences were statistically significant for most comparisons (p < 0.0001), with the exception of selected 4- to 5-year AUPRC comparisons and the 5-year AUROC comparison in SHZS.

### Performance comparison between survival and PCP-HF Models

As shown in **Table 2**, the survival model substantially improved risk reclassification relative to PCP-HF across the 1- to 5-year horizons. The categorical NRI remained consistently positive, decreasing modestly from 0.61 (95% CI, 0.59–0.62) at 1 year to 0.52 (95% CI, 0.50–0.54) at 5 years, while the continuous NRI declined from 0.92 (95% CI, 0.89–0.95) to 0.86 (95% CI, 0.83–0.89). In contrast, IDI increased from 0.18 (95% CI, 0.17–0.19) at 1 year to 0.22 (95% CI, 0.21–0.23) at 5 years, indicating improved separation between predicted risks for events and nonevents.

**Table 2.** Integrated Discrimination Improvement and Net Reclassification Improvement of the Survival Model Compared With the PCP-HF Score.

| Metrics | Year 1 | Year 2 | Year 3 | Year 4 | Year 5 |
| --- | --- | --- | --- | --- | --- |
| Categorical NRI | 0.61 (0.59-0.62) | 0.59 (0.57-0.60) | 0.57 (0.55-0.59) | 0.54 (0.52-0.56) | 0.52 (0.50-0.54) |
| Continuous NRI | 0.92 (0.89-0.95) | 0.88 (0.86-0.91) | 0.87 (0.84-0.90) | 0.87 (0.84-0.90) | 0.86 (0.83-0.89) |
| IDI | 0.18 (0.17-0.19) | 0.19 (0.18-0.20) | 0.20 (0.19-0.21) | 0.21 (0.20-0.22) | 0.22 (0.21-0.23) |

Across all prediction horizons, the survival model also outperformed PCP-HF in discrimination, precision–recall performance, with consistently lower Brier scores and higher net benefit on decision curve analysis (**Figure S6**).

### Biological exploration

To investigate structural associations underlying AI-ECG predictions, we correlated AI-ECG predictions with echocardiographic parameters in the BIDMC cohort. The analysis indicated that AI-ECG outputs were related to markers of cardiac chamber remodeling. The strongest correlations were observed with left ventricular systolic and structural indices, including left ventricular ejection fraction, left ventricular end-systolic diameter, and left ventricular end-diastolic diameter. AI-ECG predictions showed positive associations with left ventricular end-systolic and end-diastolic diameters and a negative association with left ventricular ejection fraction (**Figure 5**).

**Figure 5.**
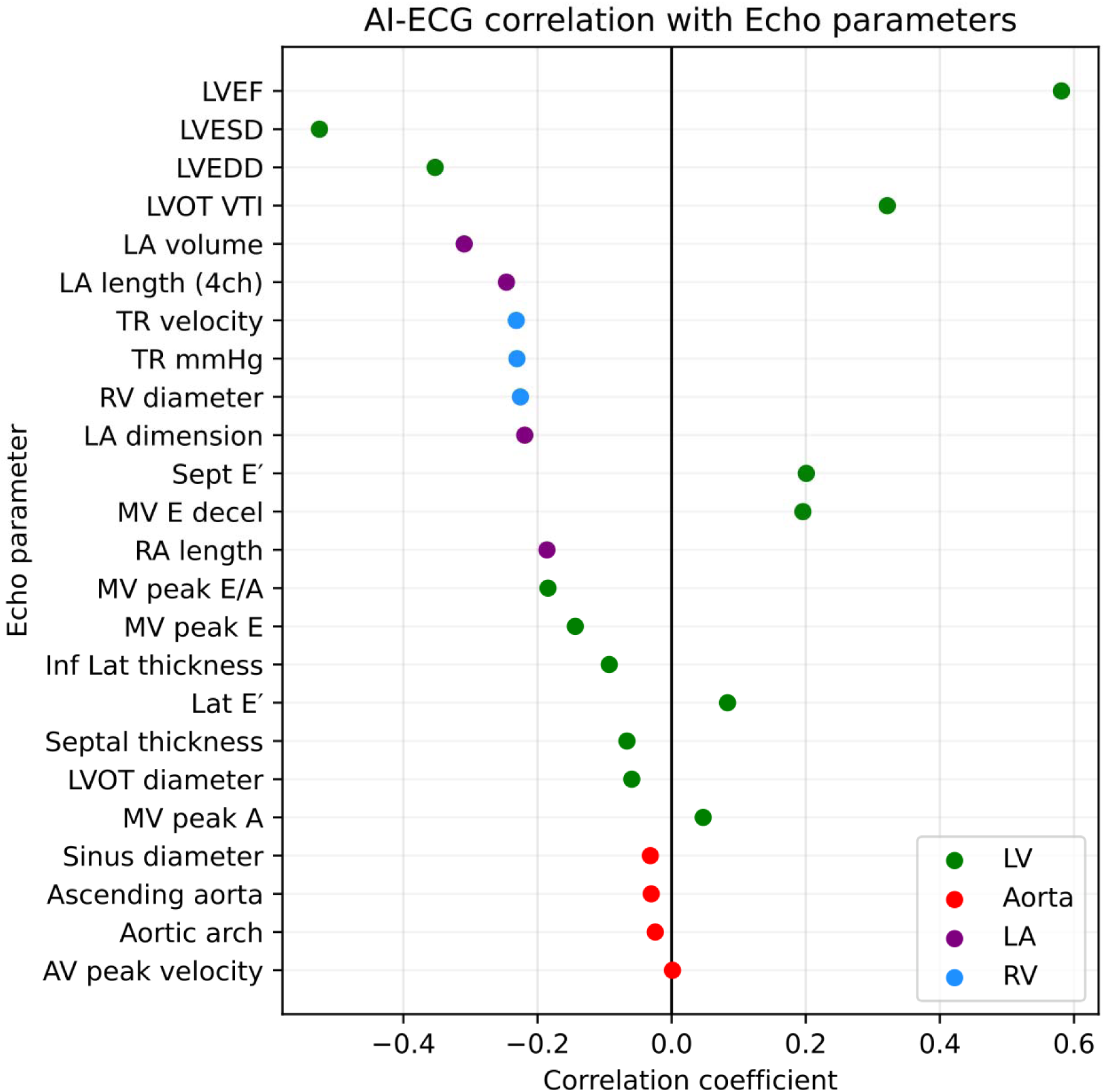
Correlation between AI-ECG representations and echocardiographic parameters. The x-axis shows correlation coefficients, and parameters are grouped by cardiac structures (LV, LA, RV, and aorta). Positive and negative values indicate direct and inverse associations, respectively.

### ECG morphologies associated with model predictions

**Figures 6 and 7** summarize the interpretability analyses of the model’s ECG representations. SHAP analysis identified several latent dimensions (z4, z47, z68, z43, and z53) as the most influential contributors to predicted HFrEF risk (**Figure 6A**). Latent-space traversal demonstrated that systematic variation of these dimensions produced coherent and physiologically plausible changes in ECG morphology across multiple leads, particularly affecting QRS amplitude, QRS duration, and repolarization features **(Figure 6B**). Correlation analyses further showed consistent associations between these latent features and conventional ECG parameters—including QRS duration, QTc interval, and heart rate-in both the SHZS and BIDMC cohorts (**Figure 6C** and **6D**).

**Figure 6.**
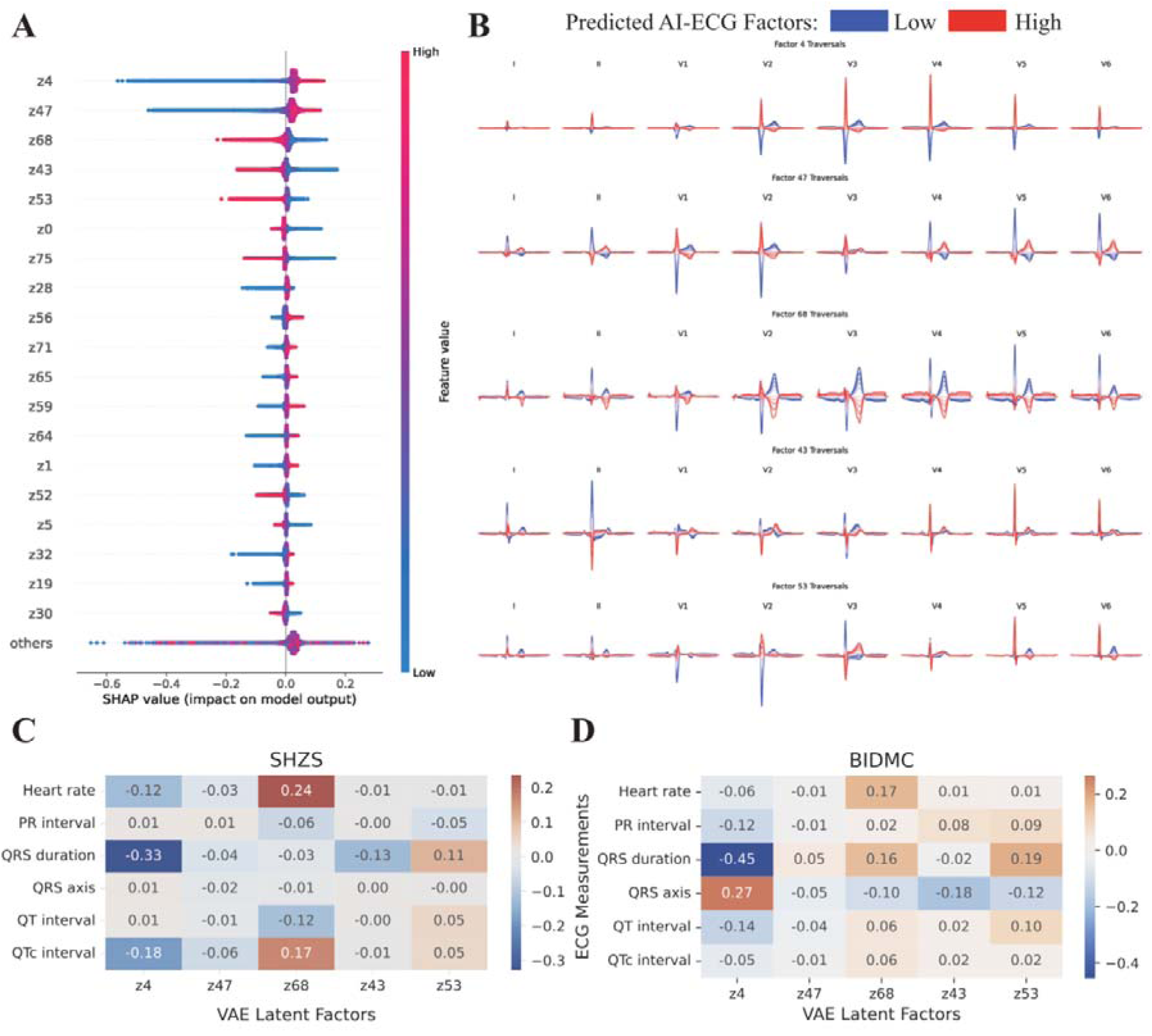
Interpretability analysis of AI-ECG latent representations. (A) SHAP summary plot showing the contribution of key VAE latent factors to model predictions. (B) Latent factor traversals illustrating ECG waveform changes across leads for representative factors at low and high values. (C-D) Heatmaps showing correlations between selected latent factors and conventional ECG measurements in the SHZS and BIDMC cohorts, respectively.

**Figure 7.**
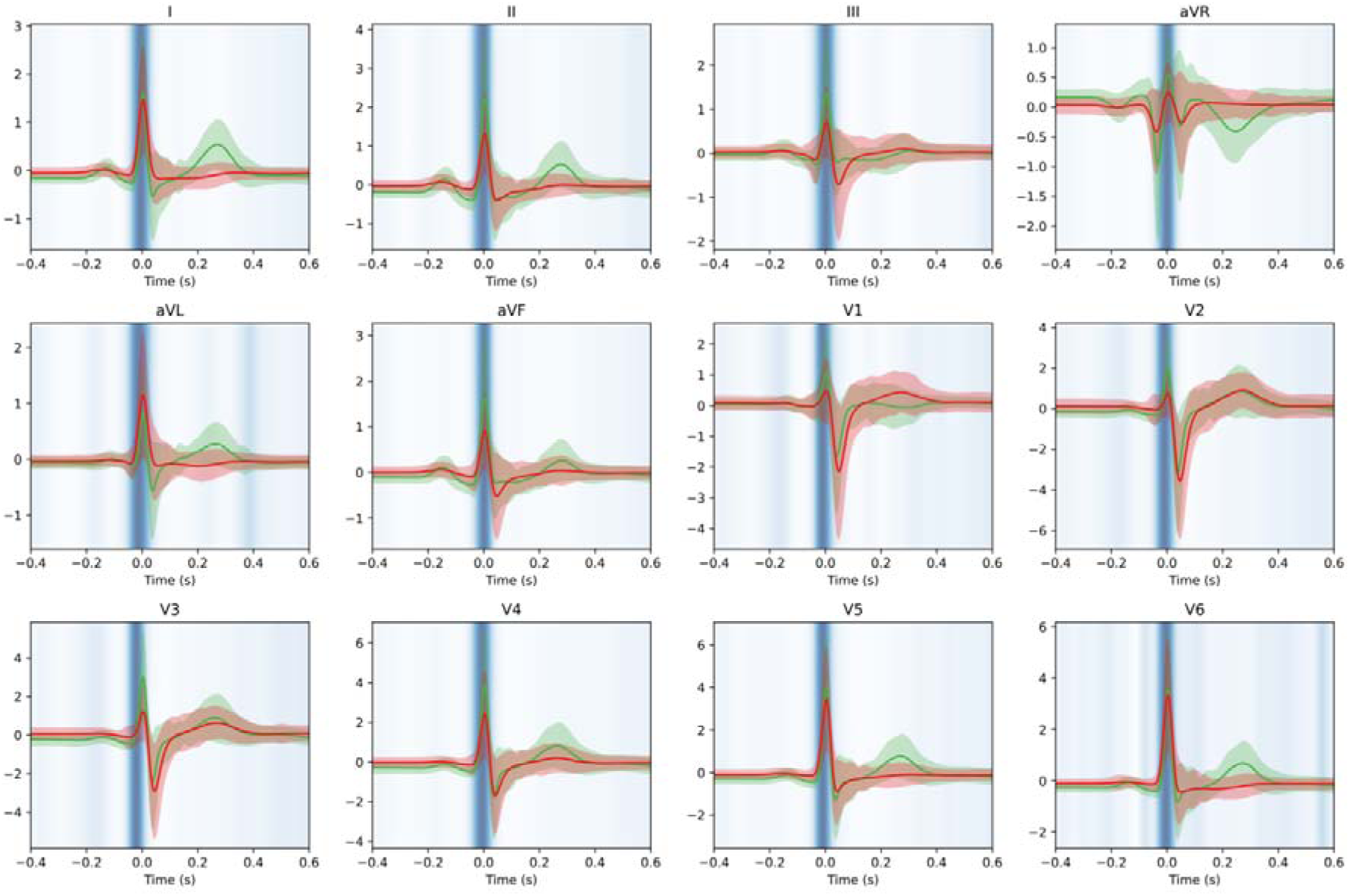
Model attention and waveform differences across AI-ECG risk groups. Median-beat ECG waveforms across all 12 leads are shown for the low-risk (green) and high-risk (red) groups, with shaded bands representing +/-1 standard deviation within each group. Overlaid Grad-CAM heatmaps identify temporal regions contributing most strongly to model predictions, with darker blue denoting greater model attention.

Median-beat reconstructions combined with Grad-CAM attribution mapping provided complementary evidence by identifying the cardiac-cycle segments that most strongly influenced model predictions (**Figure 7**). High-risk patients exhibited marked alterations in QRS morphology-including changes in amplitude, duration, and contour-along with prominent ST-T abnormalities, characterized by flattened or blunted T-wave amplitudes across multiple leads. These activation patterns were reproducible across limb and precordial leads, indicating consistent model focus on regions reflecting ventricular depolarization and repolarization dynamics.

## Discussion

In this large multicenter study, we developed deep learning models based on raw ECG data for time-to-event prediction of HFrEF. Using ECG-TTE pairs from Chinese and U.S. cohorts, we established a survival-based framework that generated individualized probabilities of incident HFrEF across follow-up. Model performance was comprehensively evaluated using complementary metrics assessing discrimination, calibration, precision–recall characteristics, and clinical utility, with consistent results across age, sex, and race subgroups. We further applied two complementary interpretability approaches that linked model predictions to latent electrophysiological features and ECG waveform regions relevant to ventricular dysfunction. Together, these findings support AI-ECG as a scalable and clinically interpretable approach for earlier HFrEF risk stratification across diverse clinical settings.

### Advancing from static to time-to-event HF risk prediction

Most existing heart failure prediction models focus on diagnostic tasks or prognostic approaches that estimate risk within predefined follow-up intervals**^[2, 19]^**. Although recent AI-ECG models have expanded prognostic capability by predicting future HF risk, these approaches still rely on static representations of risk and do not capture how risk evolves over time or identify when patients transition into periods of heightened vulnerability.

Time-to-event risk modeling addresses this limitation by estimating individualized risk trajectories rather than single-point predictions, thereby shifting HF prediction from categorical risk labeling toward a longitudinal understanding of disease progression. Such an approach aligns more closely with the chronic and progressive nature of HF and provides a framework for identifying when risk intensifies, not merely whether risk is present. By quantifying changes in predicted risk across follow-up, survival-based models may help anticipate transitions from compensated to vulnerable states and support earlier clinical reassessment or closer longitudinal monitoring.

In clinical practice, the timing of risk escalation is often more actionable than the mere presence of elevated risk. Guideline-directed medical therapy (GDMT) for HFrEF confers the greatest benefit when initiated early, before irreversible ventricular remodeling and repeated decompensation occur**^[3,5,6]^**. Likewise, early identification of patients approaching symptomatic deterioration may allow intensified surveillance, timely imaging reassessment, or optimization of pharmacologic therapy before first hospitalization, an event strongly associated with subsequent mortality and healthcare burden**^[1]^**. This may be particularly relevant for individuals with asymptomatic left ventricular systolic dysfunction or high-risk comorbid profiles, in whom clinical deterioration may occur silently and unpredictably.

### Comprehensive assessment of model performance

Evaluating model performance should not be confined to any single metric, as relying on measures such as AUC alone may obscure the true behavior of a predictive system, given that AUC is highly sensitive to population characteristics and disease prevalence.**^[24]^** Therefore, we adopted a comprehensive evaluation strategy to provide a more rigorous and balanced assessment of the model’s strengths and limitations.

Across all three cohorts, our model demonstrated uniformly strong and consistent performance. The combination of the C-index and time-dependent AUROC showed steady and high discrimination across 1-5 years in both internal and external cohorts. AUPRC values remained high despite the low incidence of events, supporting the model’s robustness under class imbalance. The Brier scores increased only modestly over time and remained consistently low across all cohorts, while the calibration curves displayed close alignment between predicted and observed risks. Taken together, these findings confirm reliable probability estimation throughout the 1to 5-year prediction window. Additionally, DCA highlighted the model’s clinical relevance, demonstrating meaningful net benefit across a broad range of threshold probabilities, with the most pronounced advantage observed during the early and mid-term periods. These complementary evaluation metrics emphasize not only the statistical rigor of the model but also its practical utility, reinforcing its potential applicability in real-world HF management.

### Comparison with established risk scores

The performance differences observed between the survival model and PCP-HF likely reflect important distinctions in model structure and clinical application. PCP-HF was developed as a conventional regression-based risk score for population-level screening using a limited set of static baseline variables and has shown value in broad community populations**.^[25, 26]^** By contrast, our survival framework was designed to estimate individualized risk over time by directly modeling time-to-event information and accounting for censoring, thereby providing a more granular characterization of longitudinal risk.

Among established heart failure risk scores, several widely used models including the ARIC HF Risk Score and PREVENT-HF require clinical, laboratory, or longitudinal variables that were not uniformly available across all study cohorts. **^[27]^** PCP-HF was therefore selected as the reference model because its required features could be reliably derived within the BIDMC cohort, enabling a fair and methodologically consistent comparison. This also highlights a practical advantage of ECG-based risk modeling, as conventional scores often depend on broader baseline phenotyping, whereas our survival framework generates individualized HF risk estimates from ECG data alone. Such simplicity may improve scalability and support implementation in routine clinical settings where more extensive phenotypic data is unavailable.

The observed improvements in discrimination, calibration, reclassification, and decision-curve performance suggest that time-to-event modeling may refine risk stratification beyond conventional static risk scores. Rather than replacing established tools, this approach may complement them by providing more individualized and temporally informative risk assessments, particularly in clinical settings that prioritize longitudinal monitoring and personalized management.

### Biological basis for AI-ECG prediction of HFrEF progression

The biological basis underlying the ability of the AI-ECG model to predict future systolic dysfunction likely reflects its sensitivity to early myocardial electrical alterations associated with subclinical cardiac remodeling. In the present study, this remodeling process was examined through correlations between AI-ECG-derived risk scores and baseline echocardiographic parameters, as illustrated in **Figure 5**. The observed associations provide insight into the pathophysiological substrates captured by the model prior to overt deterioration in left ventricular systolic function.

Notably, AI-ECG predictions demonstrated the strongest correlations with echocardiographic indices of left ventricular structure and function, including LVEF, LV end-diastolic and end-systolic dimensions, and LV outflow tract velocity-time integral. These findings suggest that the model is primarily detecting electrical signatures related to early LV dilation, impaired contractility, and altered ventricular mechanics, which are hallmarks of the remodeling process that precedes clinically overt heart failure. In parallel, consistent associations with left atrial volume and left atrial length further indicate sensitivity to chronic elevation of LV filling pressures and atrial remodeling, which often accompany progressive systolic dysfunction**. ^[28, 29]^**

In addition to left-sided parameters, moderate correlations with tricuspid regurgitation velocity and right ventricular dimensions were observed, suggesting that the AI-ECG signal may also capture downstream consequences of left-sided disease, including secondary pulmonary hypertension and right ventricular loading. In contrast, correlations with aortic dimensions and aortic valve velocities were comparatively weaker, consistent with the possibility that the predictive capability of the model may be driven predominantly by myocardial and chamber-level remodeling rather than isolated vascular or valvular structural changes.

These findings support a biologically plausible mechanism whereby the AI-ECG model identifies early electrical manifestations of myocardial remodeling across multiple cardiac chambers. By capturing subclinical alterations in ventricular function, atrial size, and secondary right-sided involvement, the model is able to distinguish individuals at risk of future systolic deterioration before the onset of overt heart failure, thereby providing mechanistic support for its prognostic performance.

### Model interpretability

The integration of artificial intelligence into clinical workflows is often hindered by concerns regarding the opacity of model decision-making. **^[25]^**Therefore, strengthening transparency and demonstrating alignment with known biological mechanisms are essential for building clinician confidence and supporting responsible implementation.

In our study, the heatmap results showed strong concordance with the VAE-derived waveform analyses, reinforcing the credibility of the model’s interpretability. For example, one of the most influential latent features (Z4 in the model representation) showed a strong association with conventional ECG parameters and was inversely related to QRS duration (**Figure 6C**).This relationship was mirrored in the latent-space traversal, where patients with higher predicted HF risk displayed visibly wider QRS complexes (**Figure 6B**). Notably, these patterns were consistent with the findings from median-beat reconstructions combined with Grad-CAM, which also highlighted QRS widening as a principal contributor to model predictions. Such agreements between statistical associations and morphology-based reconstructions support the notion that the model is leveraging physiologically meaningful features rather than spurious patterns.

Moreover, the most influential latent factors corresponded to heart rate, QRS duration, and QT interval, which are electrophysiologic markers closely linked to the pathophysiology and prognosis of HFrEF (**Figure6Cand 6D**). Elevated resting heart rate reflects heightened sympathetic activation and impaired autonomic regulation and has long been recognized as a marker of worse clinical status, as evidenced by the inclusion of β-blockers in standard HFrEF therapy to control excessive heart rate.**^[30]^** Similarly, prolonged QRS duration indicates intraventricular conduction delay and mechanical dyssynchrony caused by structural remodeling and is consistently associated with increased hospitalization and mortality. **^[31]^**QT interval prolongation reflects repolarization abnormalities related to fibrosis and neurohormonal activation and is associated with heightened arrhythmic risk. **^[32]^**In addition to these features, the Grad-CAM visualizations also identified flattened T waves and low QRS voltage, which represent characteristic morphologic abnormalities frequently observed in myocardial ischemia and cardiomyopathies such as cardiac amyloidosis**.^[33]^**

The clinical relevance of these ECG features supports the biological plausibility of interpretability findings, demonstrating that the model is capturing electrophysiologic signals that align with established mechanisms and risk pathways in HFrEF.

### Generalizability across populations and settings

Our model demonstrated consistently strong performance across three independent cohorts from China and the United States, with stable results across age, sex, and race subgroups and across multiple evaluation domains, including discrimination, calibration, prediction error, and clinical utility. This consistency across heterogeneous populations supports the robustness and generalizability of the framework, suggesting that its predictive performance is not restricted to a specific demographic group or healthcare setting.

An additional strength of the model is its reliance on standard ECG data as the sole input, which is routinely available in clinical practice and substantially more accessible than advanced imaging modalities such as echocardiography or cardiac magnetic resonance imaging.This simplicity may facilitate implementation across diverse clinical environments where ECGs are routinely obtained. By enabling earlier identification of patients at risk of HFrEF, the model may support more timely risk stratification, better targeting of follow-up evaluation, and earlier access to evidence-based therapies and device-based interventions.

## Limitation

First, several established heart failure risk scores require clinical, laboratory, or longitudinal variables that were not uniformly captured across our three cohorts. As a result, PCP-HF was selected as the reference model for comparison and could be evaluated only within the BIDMC cohort, where all required inputs were available. This constraint also reflects the practical challenges of applying traditional risk scores in real-world settings, where key variables are frequently missing. Second, although the AI-ECG model effectively identifies individuals at elevated risk of incident heart failure, it remains uncertain whether the risk captured by the model is modifiable through medical or lifestyle intervention. Whether early clinical actions guided by AI-ECG predictions can improve outcomes requires further study. Third, this study is retrospective, and residual confounding may remain despite multi-center external validation. Prospective investigations, ideally randomized controlled trials, are needed to determine whether incorporating AI-ECG–based risk stratification into clinical care improves patient management and long-term outcomes.

## Conclusion

In this multinational study, we showed that a deep learning model trained on raw ECG data can generate individualized time-to-event estimates of future HFrEF risk with robust and consistent performance across diverse populations. The model demonstrated strong discrimination, reliable calibration, and meaningful clinical utility, supported by interpretability analyses with biological plausibility linking ECG-derived signals to subclinical cardiac remodeling. Although prospective studies are needed to determine whether AI-ECG–guided risk estimation can directly inform clinical management or improve outcomes, our findings highlight the potential of AI-enabled ECG analysis as a scalable and accessible approach for earlier HFrEF risk stratification within routine clinical workflows.

## Supporting information

none.

## Data Availability

All data produced in the present study are not available to public due to ethical requirements.

## Acknowledgements

The authors thank all participating hospitals for their collaboration and technical assistance.

## Clinical trial number

ClinicalTrials.gov (NCT07519434)

## Notes

### Competing Interest Statement

The authors have declared no competing interest.

### Author Declarations

The study was approved by the Institutional Research Board of Zhongshan Hospital (No. B2023-253R) with a waiver of patient consent.

