## Supplementary material for "Electrocardiogram-Based Deep Learning for Time-Resolved Prediction of Heart Failure With Reduced Ejection Fraction: A Multinational Study": none.

**’ by Lei Pan, et al.**

**FigureS1 Time-dependent calibration performance of the AI-ECG survival model across cohorts**

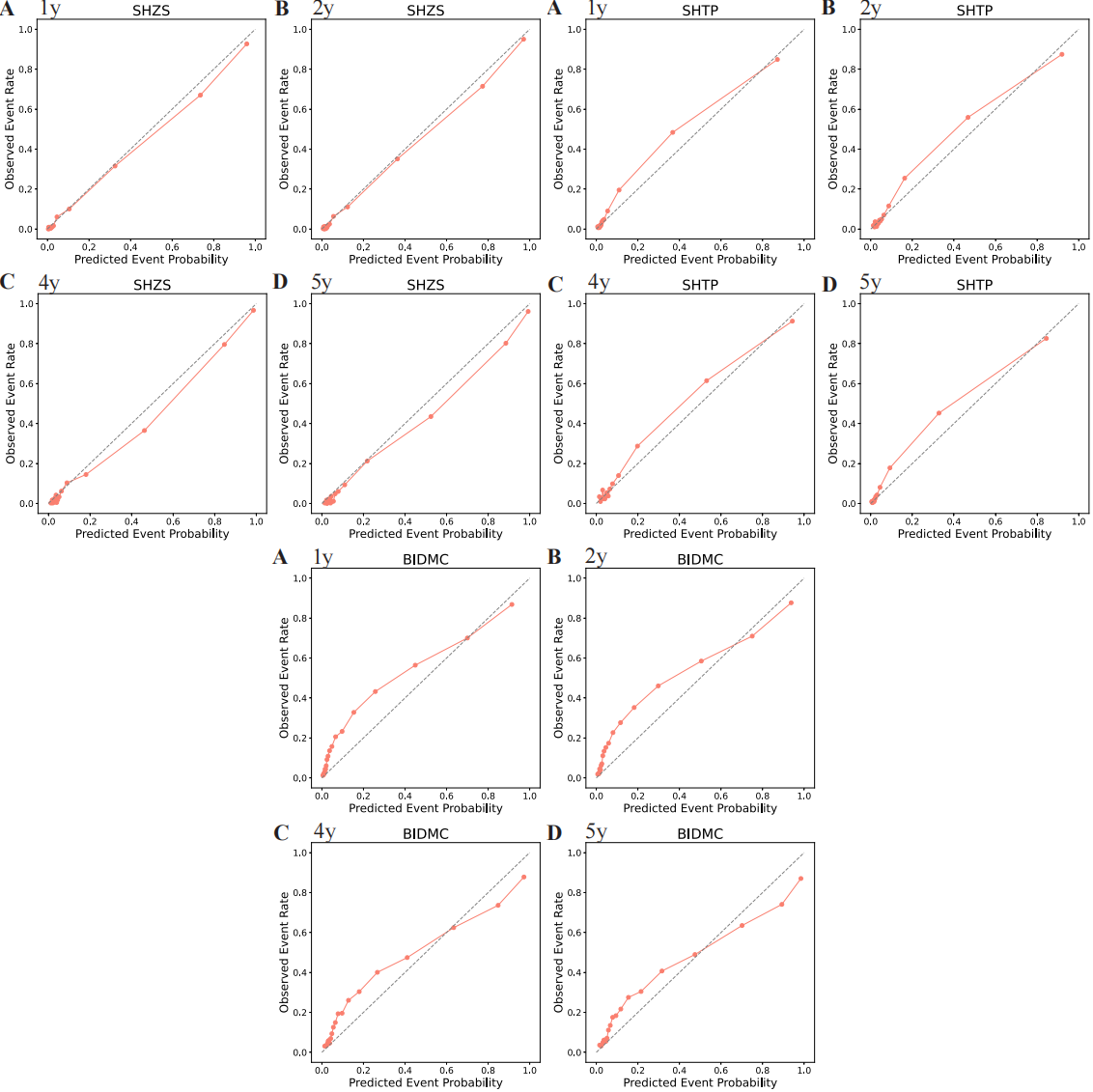

**FigureS2 Time-dependent decision curve analysis across cohorts**

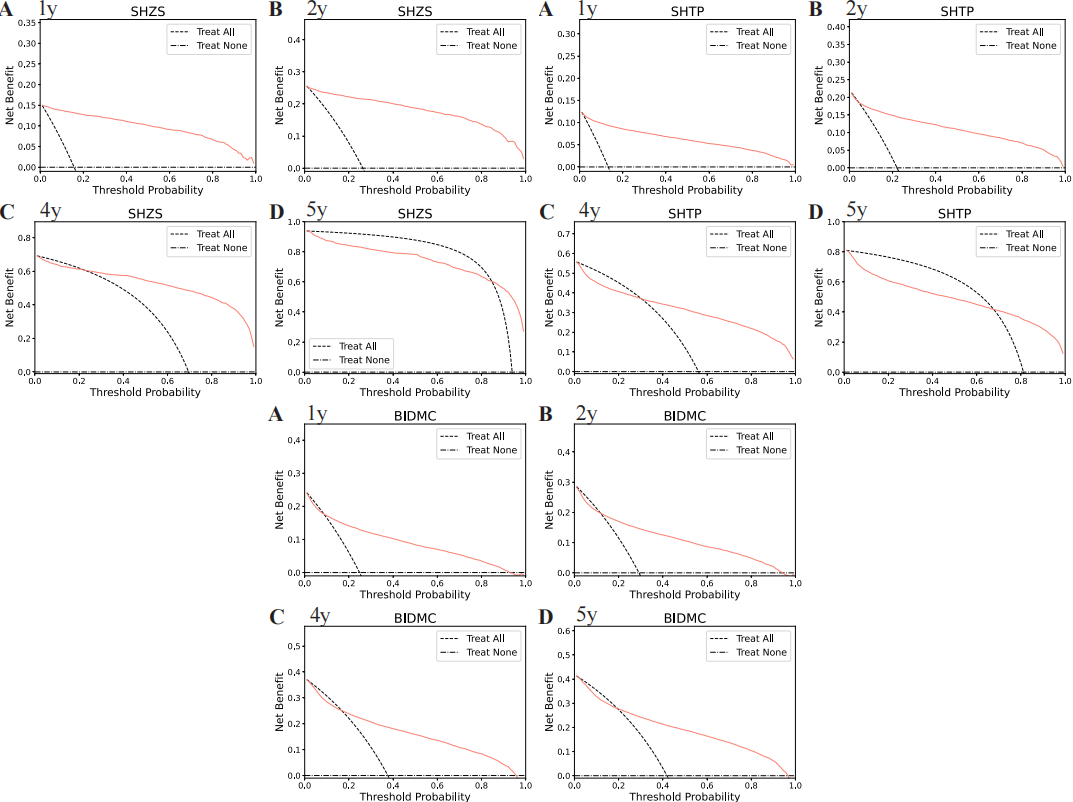

**FigureS3 Kaplan–Meier survival curves stratified by AI-ECG–predicted risk across cohorts and prediction horizons**

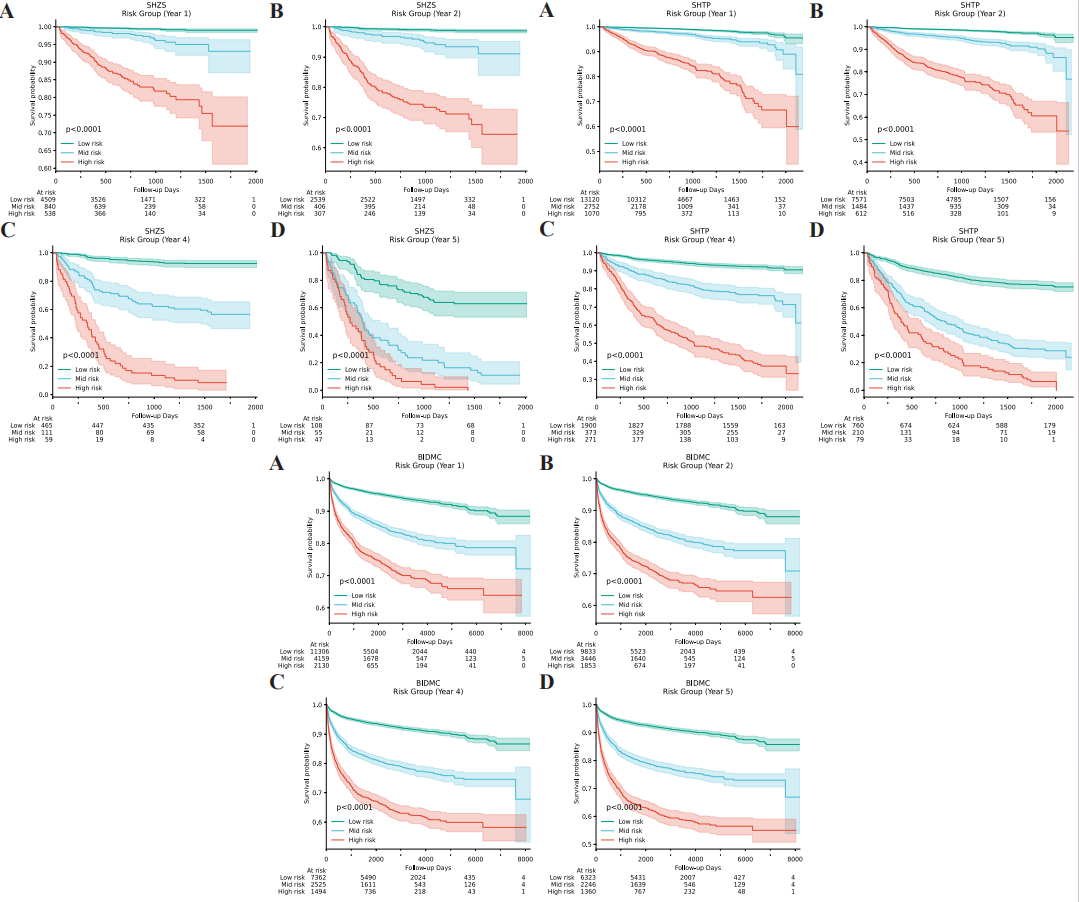

**FigureS4 Model performance across demographic subgroups**

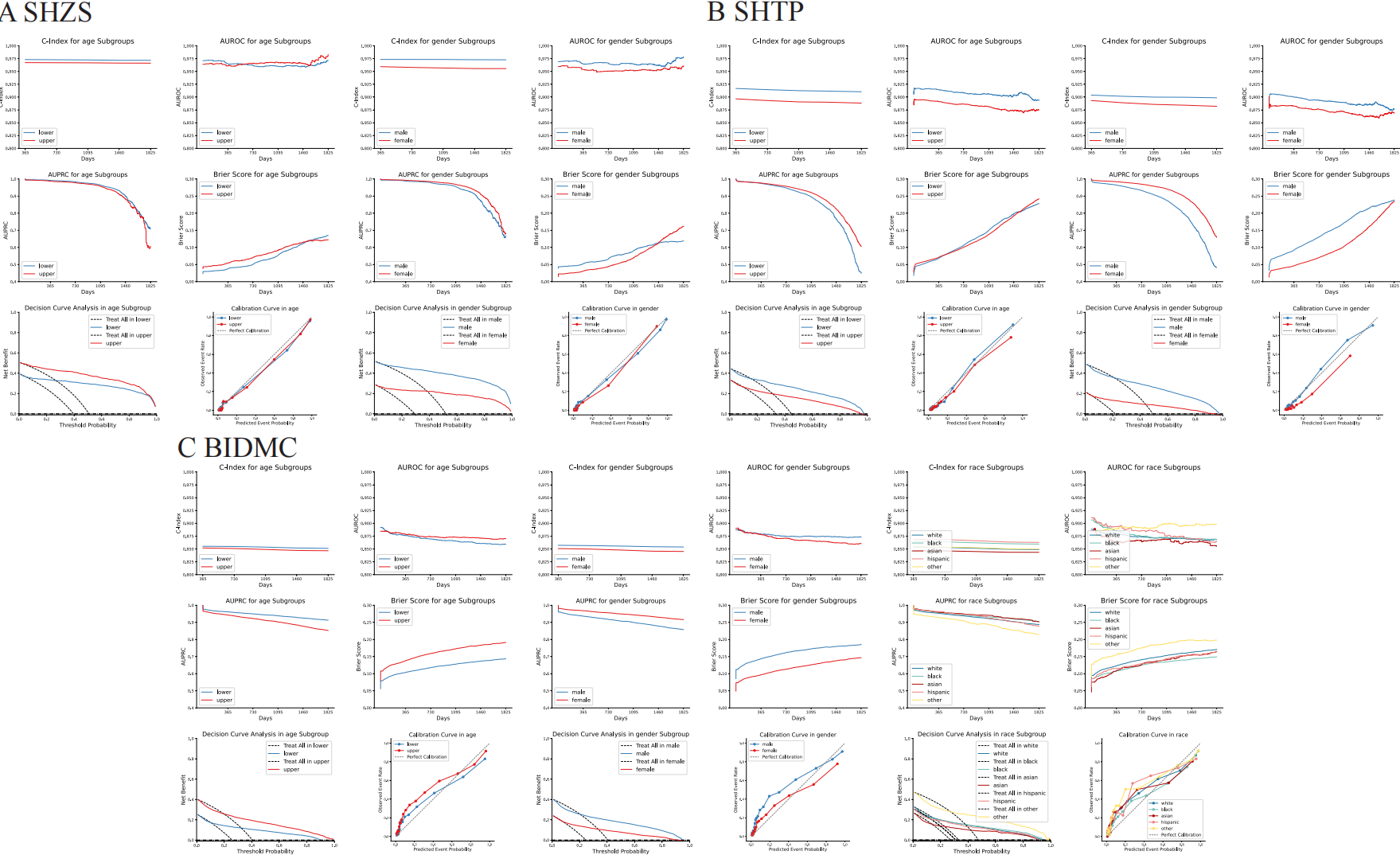

**FigureS5 Comparison of survival-based and classification-based AI-ECG models across cohorts**

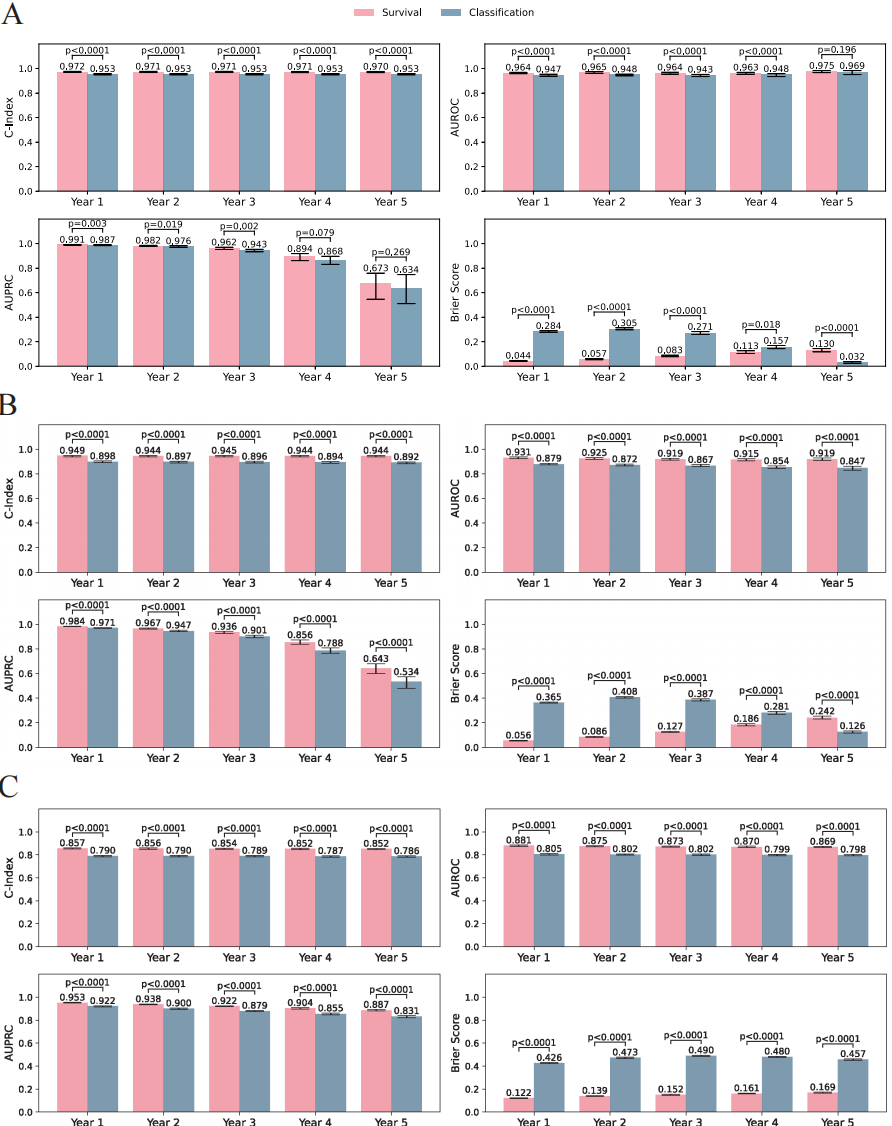

**FigureS6 Comparative predictive performance and clinical utility of the AI-ECG survival model and PCP-HF in the BIDMC cohort**

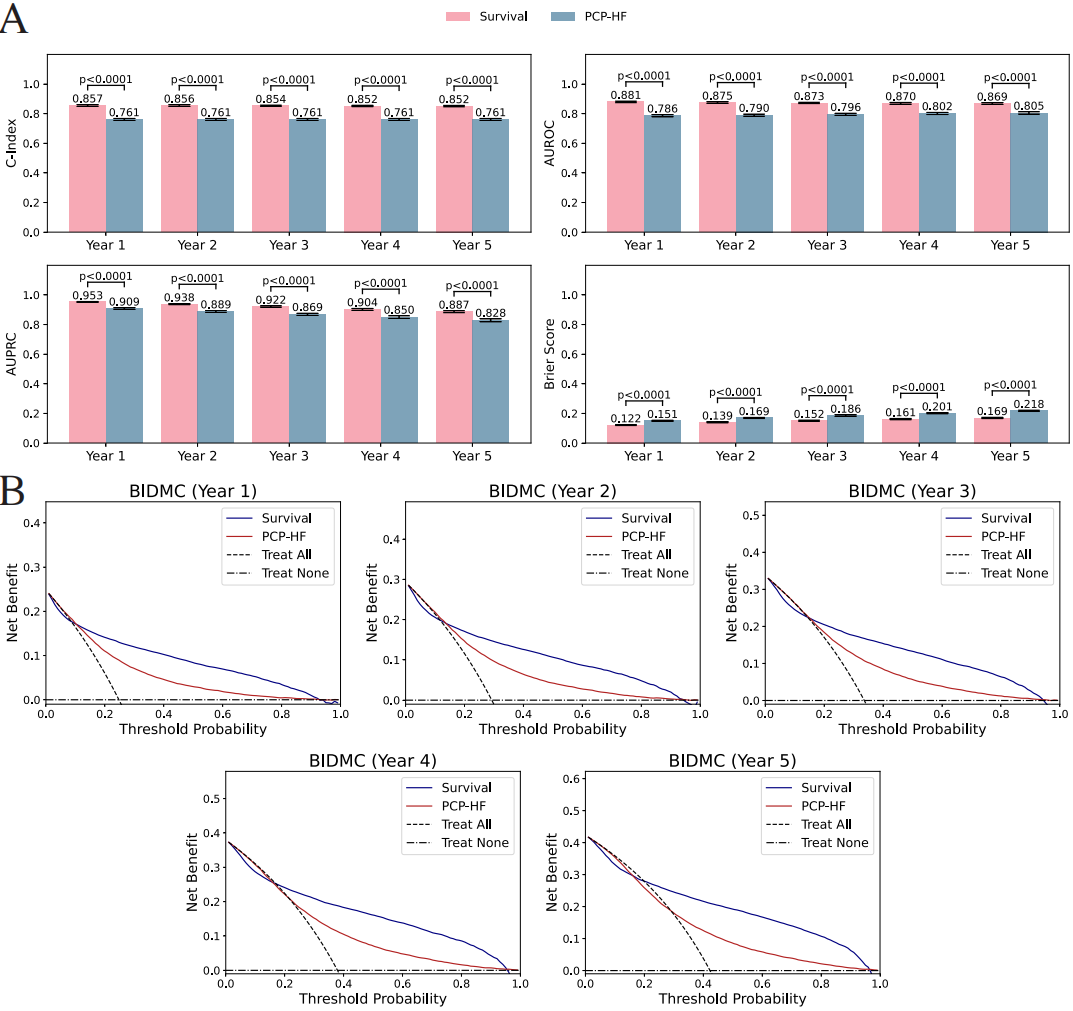
